# Identifying Prediabetes in Canadian Populations Using Machine Learning

**DOI:** 10.1101/2024.02.03.24302301

**Authors:** Katherine Lu, Paijani Sheth, Zhi Lin Zhou, Kamyar Kazari, Aziz Guergachi, Karim Keshavjee, Mohammad Noaeen, Zahra Shakeri

**Author notes:** These authors contributed equally to this work and share first authorship (Katherine Lu, Paijani Sheth, Zhi Lin Zhou).

## Abstract

Prediabetes is a critical health condition characterized by elevated blood glucose levels that fall below the threshold for Type 2 diabetes (T2D) diagnosis. Accurate identification of prediabetes is essential to forestall the progression to T2D among at-risk individuals. This study aims to pinpoint the most effective machine learning (ML) model for prediabetes prediction and to elucidate the key biological variables critical for distinguishing individuals with prediabetes. Utilizing data from the Canadian Primary Care Sentinel Surveillance Network (CPCSSN), our analysis included 6,414 participants identified as either nondiabetic or prediabetic. A rigorous selection process led to the identification of ten variables for the study, informed by literature review, data completeness, and the evaluation of collinearity. Our comparative analysis of seven ML models revealed that the Deep Neural Network (DNN), enhanced with early stop regularization, outshined others by achieving a recall rate of 60%. This model’s performance underscores its potential in effectively identifying prediabetic individuals, showcasing the strategic integration of ML in healthcare. While the model reflects a significant advancement in prediabetes prediction, it also opens avenues for further research to refine prediction accuracy, possibly by integrating novel biological markers or exploring alternative modeling techniques. The results of our work represent a pivotal step forward in the early detection of prediabetes, contributing significantly to preventive healthcare measures and the broader fight against the global epidemic of Type 2 diabetes.

## I. Introduction

Type 2 diabetes (T2D) is a detrimental condition that affects millions of people worldwide [1]. The precursor to T2D is prediabetes, a condition where blood glucose levels are elevated but below the diagnosis threshold for T2D [2]. Prediabetes is a common but reversible condition; according to an American Diabetes Association panel, up to 70% of individuals with prediabetes will develop diabetes [3]. Therefore, it is imperative to address prediabetes during the onset to prevent it from developing into T2D in the future.

Prediabetes is specifically defined by a fasting blood sugar (FBS) level between 6.1 and 6.9 mmol/L [4]. Many unhealthy lifestyle factors contribute to the development of T2D, so it is important to implement preventative measures such as exercise programs when individuals reach the pre-diabetes stage. Prediabetes also shares many of the same indicators as T2D. For example, hypertension [5], high total cholesterol [6], depression [7], glucocorticoids [8], chronic obstructive pulmonary disease (COPD) [9], osteoarthritis [10], and increased age have all been found to be associated with greater risk of developing T2D. Because risk factors of diabetes are well-known, many studies have applied machine learning (ML) models to clinical data to predict diabetes [11–16]. However, there are no Canadian studies developing prescriptive ML models to predict prediabetes. The onset of prediabetes is insidious, and most prediabetic individuals are unaware of their condition [17]. Therefore, accurately detecting prediabetes with ML models will improve health outcomes and reduce the burden on the healthcare system. Our study aims to identify the best ML model to predict prediabetes, and determine the important clvariables in pre-diabetic individuals.

Building on the foundation laid by previous research, This study takes a novel approach by creating machine learning models specifically designed for the Canadian population to predict prediabetes. By identifying key variables and leveraging cutting-edge predictive analytics, we aim to advance early detection methods, ultimately fostering preventative measures against the transition from prediabetes to Type 2 diabetes. This effort not only aims to enhance awareness and management of prediabetes but also sets a precedent for utilizing technology-driven approaches in public health strategies, marking a significant step towards mitigating the global challenge of diabetes.

## II. Methods

### A. Data Source and Study Population

Our study uses data from the Canadian Primary Care Sentinel Surveillance Network (CPCSSN) [18]. Briefly, the CPCSSN compiles de-identified electronic medical record (EMR) data from 14 participating primary care networks across Canada. CPCSSN data is typically extracted from EMRs twice per year, and only structured data (e.g., not physician notes) is extracted. Details of CPCSSN data are described elsewhere [18, 19].

The data for our study is a random sample (n=10,000) of a CPCSSN subset that is comprised of patient records that precede the onset of diabetes. We restricted our study cohort to nondiabetic and prediabetic patients resulting in a final sample size of 6,414. Diabetic patients were excluded if they either had a fasting blood sugar (FBS) greater than 6.9 or an A1c level over 6.4% [4].

### B. Data Processing and Feature Engineering

We selected a total of 10 variables based on existing literature supporting an association with diabetes [5–10]. We removed variables with over 50% missing observations. If variables were collinear, the most appropriate variable was selected; for example, total cholesterol includes low-density lipoprotein (LDL) cholesterol and high-density lipoprotein (HDL) cholesterol, so we kept total cholesterol and removed LDL and HDL. The final variables we included in our analysis were: age, body mass index (BMI), total cholesterol, depression status, hypertension status, presence of osteoporosis, chronic obstructive pulmonary disease (COPD), corticosteroid use, hypertension medication use (HMU), and sex. Corticosteroid and HMU were recategorized from a descriptive variable with all medication names to a binary variable of whether patients used any hypertension medications. The distributions, correlations, and missingness of all selected variables were explored. The class balance of the target, prediabetes, was also explored.

The outcome variable, presence of prediabetes, was created based on both FBS and A1c values. Patients were labeled as prediabetic if they had an FBS score between 6.1–6.9 mmol/L, or an A1c between 6.0–6.4% [4].

### C. Prediction Models Development

We selected seven machine learning models to predict for prediabetes: logistic regression, Random Forest, XGBoost (extreme gradient boosting), mixed Naive Bayes, KNN (K-Nearest Neighbours), SVM (support vector machine), and Deep Neural Network (DNN). These models were chosen to explore model types with different strengths such as computational efficiency and robustness to overfitting, and for their proven performance in diagnosing prediabetes or T2D in previous studies [12, 20–23]. The data engineering process for all models involved splitting the data into training (75%), validation (12.5%) and testing (12.5%) sets. Following splitting, all data sets were imputed for missing values using the median. Median imputation, chosen for its preservation of the total cholesterol distribution and outlier resilience, was applied to the sole variable with missing values. Post-imputation, all datasets underwent normalization.

The DNN model’s architecture involved defining aspects of the neuron and hidden layers. Before training, the data was stored into PyTorch tensors to improve efficiency during the forward pass and back propagation processes. The model was run on the training set, and the Binary Cross-Entropy Loss function between the predictions and true value was computed and visualized as loss over epochs. This was important in determining whether the model was learning over epochs. To reduce risk of overfitting, three DNN models with different types of regularization techniques were explored: L2 regularization, drop out, and early stopping. Hyperparameters were tuned manually. The highest performing parameters were: learning rate of 0.001, batch size of 25, 10 hidden units, and drop out probability of 0.5. Using the optimal hyperparameters, model performance was assessed on the validation set using all three regularization techniques and the best performing model was finally run on the test dataset.

Lastly, to identify the important variables in predicting prediabetes, a SHAP (SHapley Additive exPlanations) analysis was conducted on Google Colab and a SHAP dot plot was displayed. To ensure replicability and facilitate further research, the source code of all the presented machine learning models is available on GitHub^1^.

## III. Results and Discussion

Patient demographics are illustrated in Table I. Among all variables selected, only total cholesterol had missing data. Our sample had 59% nondiabetic patients and 41% prediabetic patients, indicating no issue of class imbalance. All continuous variables (age, BMI, total cholesterol) demonstrated normal distributions. For prediabetic patients, age and BMI were slightly higher while total cholesterol was slightly lower compared to nondiabetic patients. A correlation matrix of the continuous variables showed that age and BMI had slight correlations with prediabetes (correlation coefficient around 0.1-0.4), and no correlation for total cholesterol.

**TABLE I:**
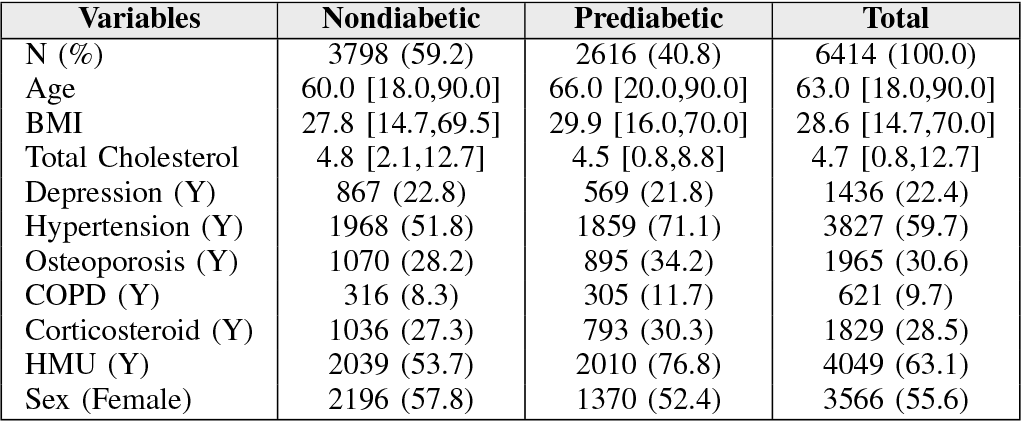
CPCSSN subset patient characteristics stratified by prediabetes status. Results are presented as median [min, max] or n (%).

### A. Prediction Models Results

Final model evaluation metrics are presented in Table II. For model evaluation, we produced and compared the following scores: accuracy, precision, recall, F1-score, and AUC. However, recall values were primarily used when assessing the performance because our goal is to minimize the number of false negatives in our study. The DNN model performed the best with a recall value of 60%, followed by logistic regression (57%), Random Forest (52%), XGBoost (51%), KNN (38%) and Naive Bayes (23%).

**TABLE II:** Performance metrics for each type of machine learning model using test sets for predicting prediabetes.

| Models | TP | FP | FN | TN | Acc | Precision | Recall | F1 | AUC |
| --- | --- | --- | --- | --- | --- | --- | --- | --- | --- |
| LR | 215 | 308 | 164 | 596 | 0.63 | 0.41 | 0.57 | 0.48 | 0.61 |
| RF | 350 | 101 | 324 | 829 | **0.74** | **0.78** | 0.52 | 0.62 | 0.71 |
| XGBoost | 272 | 190 | 265 | 556 | 0.65 | 0.59 | 0.51 | 0.54 | 0.63 |
| NB | 69 | 51 | 232 | 450 | 0.65 | 0.58 | 0.23 | 0.33 | 0.56 |
| DNN | 198 | 87 | 134 | 383 | 0.72 | 0.69 | **0.60** | **0.64** | **0.76** |
| KNN | 130 | 210 | 210 | 260 | 0.49 | 0.38 | 0.38 | 0.38 | 0.66 |
| SVM | 254 | 269 | 166 | 594 | 0.66 | 0.60 | 0.49 | 0.54 | 0.63 |

Note: Bold indicates the highest value of a metric in the column.

Using the optimal hyperparameters for the DNN model, the loss over 2000 epochs plot showed a sudden decline within the first 50 epochs before reaching a plateau around 50%. We implemented 3 regularization techniques: L2 regularization, dropout regularization, and early stopping regularization. Early stop regularization yielded the highest performance, with a validation recall of 53% after 200 epochs. Furthermore, the recall improved over epochs, suggesting that the model learned. Therefore, an early stop regularization technique was used when assessing the performance on the test data. The test recall ended up being 60% and there was a slight overall increase in performance over 200 epochs. To assess the generalizability of the model and its learned state, train and test accuracies over 200 epochs were also visualized (Figure 1). Figure 2 displays the SHAP dot plot for our selected features from highest to lowest importance in predicting prediabetes.

**Fig. 1:**
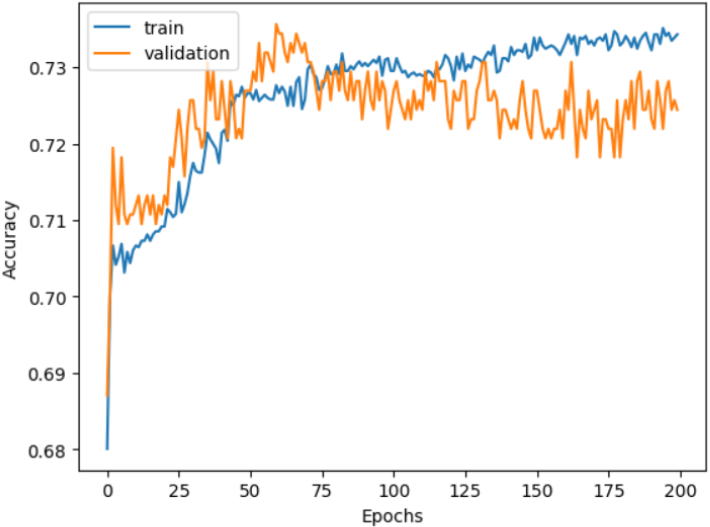
Performance plots for the DNN model for prediabetes prediction

**Fig. 2:**
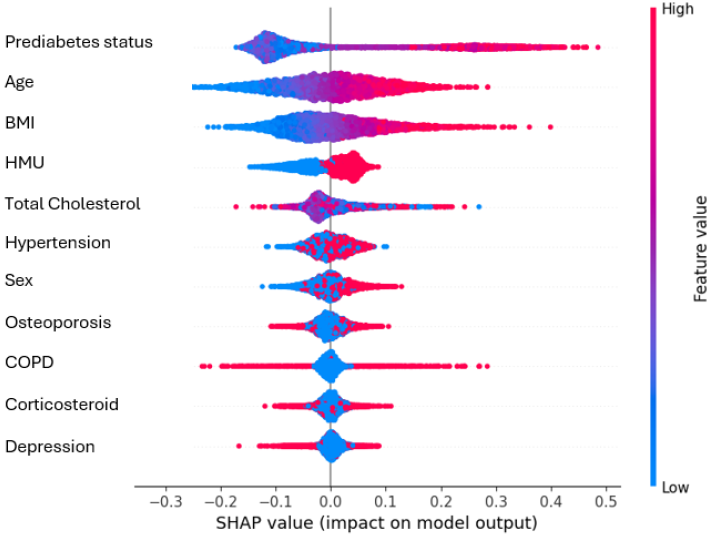
SHAP scatter plot for prediabetes prediction DNN features.

### B. Discussion

After conducting the exploratory process, it was found that the majority of the categorical features were relatively balanced when distributed among prediabetic and nondiabetic groups (Table I). Among the continuous variables, the median age and BMI of prediabetic individuals were higher compared to nondiabetic individuals. Furthermore, a correlation matrix between the continuous variables suggested no correlation among covariates as the highest correlation coefficient was around 0.2 between age and prediabetes.

The DNN model with early stop regularization had the best overall performance, having the highest recall, F1 score, and AUC (Table II). However, the primary performance metric of this research was recall, as it aimed to minimize the number of false negatives and minimize the number of prediabetic individuals left undiagnosed. Failing to identify prediabetic individuals would result in a higher risk of them developing diabetes, and interventions to manage the disease become more invasive and costly. Looking solely at accuracy, the Random Forest model appeared best performing (accuracy = 0.74), but it’s low recall of 0.52 indicated that more individuals with prediabetes were falsely diagnosed as healthy. The Neural Network model, on the other hand, exhibited an accuracy of 0.71 and a recall of 0.60 (Table II), which underscored its superior performance among the evaluated models. Despite this, we observed notable limitations in its application. Firstly, there was evidence of some overfitting to the training data, as illustrated in Figure 1. While both test and training accuracy increased and stabilized in tandem—suggesting effective learning from the training dataset—the presence of more pronounced peaks and dips in test accuracy indicates potential overfitting. This pattern suggests that the model may not generalize as effectively to unseen data. Additionally, a third limitation emerged in the form of low predictive power, evidenced by the suboptimal performance across all models. This aspect further highlights the challenges in achieving high accuracy in prediabetes prediction, underlining the need for continued refinement of machine learning approaches in this domain.

Our results are in contrast with some existing literature that use similar ML models to predict prediabetes/diabetes with better performance. For example, Lai et al. [12] obtained a recall of 73.4 using logistic regression to predict diabetes, and Choi et al. [22]obtained a recall of 74.3 using SVM to predict prediabetes . A potential reason could be that biological characteristics of nondiabetic and prediabetic individuals may overlap more compared to nondiabetic and diabetic individuals, making it more difficult to predict prediabetes. Another explanation could be that the biological features we selected were not adequate for distinguishing those with prediabetes from nondiabetics.

The results of SHAP analysis on the other hand aligns with evidence from current literature. We found that the top 3 important features in predicting prediabetes were age, BMI, and hypertension medication use. A 2016 study on characteristics associated with prediabetes and found BMI to be higher in prediabetes patients [24]. Prediabetes has been widely associated with advanced age and hypertension as well [25, 26].

The rapid expansion of ML applications for healthcare has prompted discussion regarding the unique ethical concerns of ML. For instance, if the training data for an ML model does not include different racial groups, the model may overlook biological differences that affect disease diagnosis and presentation [27]. Such oversights can introduce bias in healthcare and exacerbate health inequities. Diversity, equity, and inclusion (DEI) is an important consideration for diabetes research because thresholds and symptoms of diabetes and prediabetes can vary across populations. In Canada’s diverse population, studies have highlighted an association between lower income levels and certain racial groups with an increased diabetes risk [28, 29]. A review of relevant literature in Medline and Embase revealed a paucity of discussions on DEI within the context of machine learning applications in diabetes research. Our dataset’s exclusion of race/ethnicity information precludes DEI exploration. Enhancing DEI in diabetes ML research necessitates the inclusion of comprehensive demographic data for analysis.

## IV. Conclusion

Through identifying prediabetes, patients can take measures to reduce the chance of developing T2D and detrimental health outcomes. The goal of our study was to use machine learning models to detect prediabetes in Canadian populations by using a sample from the CPCSSN dataset. Our second objective was to determine important clinical variables for identifying prediabetic individuals. We fit seven machine learning models, with DNN showing in the best performance with a recall of 60%. The most important features in predicting prediabetes were age, BMI, and hypertension medication use, which aligns with current literature.

The DNN model’s limitation was its failure to identify approximately 40% of prediabetes cases, suggesting the current variables might not be predictive enough. This limitation stems from the restricted variable selection available in the CPCSSN dataset. Future efforts should focus on enhancing model accuracy by incorporating a wider range of variables, such as demographics, diet, physical activity, and smoking status, and considering sex-specific differences due to varying biomarker levels. An improved machine learning model for prediabetes diagnosis is essential for early detection of individuals at risk for Type 2 Diabetes, facilitating proactive preventative interventions by healthcare professionals.

## Data Availability

All data used are available online at https://cpcssn.ca/ website upon reasonable request.

## Footnotes

1 https://github.com/lilyzhizhou/Identifying-Prediabetes-ML.git

